# Resting Heart Rate as a Non-Cardiovascular Mortality Marker in Young Adults: A Population-Based Cohort Study

**DOI:** 10.64898/2026.05.20.26353745

**Authors:** Huizhen Chen, Qiu Chen, Yanzhong Wang

## Abstract

**Background:** Elevated resting heart rate (RHR) predicts mortality in older adults, primarily through cardiovascular disease (CVD). Prior cohort evidence suggests that RHR also predicts mortality in younger adults, but whether this association operates through cardiovascular or non-cardiovascular pathways has not been directly tested.

**Methods and Results:** We analyzed 3291 adults aged 20 to 49 years from NHANES 1999–2004 linked to mortality data through 2019 (median follow-up, 17.8 years; 120 deaths). RHR and heart rate reserve (HRR) were modeled per 10-bpm increment using Cox regression adjusted for demographic, lifestyle, and comorbidity covariates. Each 10-bpm RHR increase was associated with higher all-cause mortality (hazard ratio [HR], 1.26; 95% CI, 1.07–1.50; P=.007), driven by non-CVD mortality (HR, 1.28; 95% CI, 1.07–1.55; P=.009) rather than CVD mortality (HR, 1.15; 95% CI, 0.77–1.71; P=.51). A behavioral/external composite (accidents and NCHS residual causes, including suicide and liver disease) reached significance (HR, 1.35; P=.02), whereas a disease-oriented composite did not (P=.20). The association was absent before age 35 (HR, 0.98; P=.88) but pronounced at ages 35–39 (HR, 2.60; P=.001). HRR was not associated with any outcome.

**Conclusions:** In young US adults, elevated RHR predicted mortality through non-cardiovascular rather than cardiovascular pathways, concentrated among behavioral and external causes. The association emerged at age 35, below current screening thresholds. HRR under submaximal conditions carried no prognostic value. RHR in young adults may reflect global health vulnerability rather than cardiovascular risk alone.

**Clinical Perspective:** 

**What Is New?:**

- In adults aged 20 to 49 years, elevated resting heart rate predicted all-cause mortality through non-cardiovascular pathways, particularly behavioral and external causes of death, rather than cardiovascular disease.
- The resting heart rate–mortality association was absent before age 35 and emerged sharply thereafter, suggesting a risk window below the starting age of SCORE2 and the Pooled Cohort Equations.
- Heart rate reserve measured under submaximal exercise conditions showed no prognostic value across all outcomes and subgroups.

**What Are the Clinical Implications?:**

- Clinicians encountering persistently elevated resting heart rate in young adults may consider evaluating for autonomic-mediated risk factors (psychological distress, substance use, sleep disorders, chronic inflammation) rather than focusing exclusively on cardiovascular workup.
- Adults aged 35 to 49 years with elevated resting heart rate may benefit from enhanced screening not currently captured by standard cardiovascular risk assessment tools.
- Population-based surveys and primary care settings using submaximal exercise protocols should interpret heart rate reserve results with caution, as submaximal protocols may lack the physiological stress needed to unmask prognostically meaningful chronotropic impairment.

## Introduction

Elevated resting heart rate (RHR) is a well-established predictor of cardiovascular and all-cause mortality in middle-aged and older populations.^1–4^ Each 10-beat-per-minute (bpm) increase in RHR carries a 10% to 20% higher mortality risk in adults older than 50 years, with proposed mechanisms centering on sympathetic overactivation, accelerated atherosclerosis, and increased myocardial oxygen demand.^5,6^

In younger adults, the evidence is more limited and the responsible pathways less clear. The Chicago Heart Association Detection Project in Industry reported that RHR predicted noncardiovascular as well as cardiovascular mortality over 22 years, with modest associations observed in men aged 18 to 39 years at baseline.^29^ However, that analysis did not separate non-cardiovascular pathways from cardiovascular pathways in younger adults or test whether the mechanism differs by age. In adults aged 20 to 49, the atherosclerotic burden that mediates the RHR–mortality link in older populations is typically minimal,^7,20^ and the cardiovascular mechanisms invoked to explain that link may not yet be operative. If RHR does predict mortality when measured in young adulthood, the responsible pathways may differ from those in older populations. To our knowledge, no study has directly tested whether the RHR–mortality association in young adults is driven by cardiovascular or non-cardiovascular causes of death.

Heart rate reserve (HRR), defined as the difference between peak exercise heart rate and RHR, provides complementary information about cardiac chronotropic competence.^8^ Low HRR has been associated with mortality in clinical populations undergoing maximal exercise testing,^9,21^ but evidence from population-based samples using submaximal protocols is limited.

The NHANES 1999–2004 included a submaximal cardiovascular fitness treadmill examination in adults aged 12 to 49 years, with subsequent linkage to national mortality data through 2019, providing up to 20 years of follow-up. The treadmill examination yields two complementary heart rate measures: RHR, indexing resting autonomic tone, and HRR, indexing submaximal chronotropic reserve. We aimed to determine (1) whether either or both predict all-cause and cause-specific mortality in adults aged 20 to 49 years, and (2) whether any observed association operates through cardiovascular or non-cardiovascular pathways.

## Methods

### Study Design and Population

This study used data from the NHANES 1999–2000, 2001–2002, and 2003–2004 cycles, nationally representative cross-sectional surveys conducted by the National Center for Health Statistics (NCHS). The NHANES protocol was approved by the NCHS Research Ethics Review Board, and all participants provided written informed consent.

The cardiovascular fitness examination involved a submaximal treadmill test (Acuflex treadmill, Quinton Cardiology Inc) using an individualized protocol targeting 75% of age-predicted maximum heart rate. Participants aged 12 to 49 years without contraindications were eligible. We restricted the analysis to adults aged 20 to 49 years with valid resting heart rate measurements. Of 3359 participants who completed the treadmill examination, 68 (2.0%) were excluded for missing covariate data (primarily hypertension status [n = 51], BMI [n = 10], and smoking status [n = 4]), yielding an analytic sample of 3291.

### Exposures

Resting heart rate was measured as the seated, pre-exercise pulse rate recorded during the cardiovascular fitness examination. Heart rate reserve was calculated as stage 2 peak heart rate minus resting heart rate, reflecting submaximal chronotropic response. Both exposures were modeled continuously per 10-bpm increment.

### Outcomes

Mortality was ascertained through probabilistic linkage with the National Death Index through 31 December 2019, performed by the NCHS. Vital status was determined using the MORTSTAT variable. Cause of death was classified using the NCHS 10-category underlying cause-of-death variable (UCOD_LEADING): heart disease (category 1), malignant neoplasms (category 2), chronic lower respiratory disease (category 3), accidents (category 4), cerebrovascular disease (category 5), and other causes. CVD mortality was defined as heart disease or cerebrovascular disease (categories 1 and 5). Non-CVD mortality included all remaining causes.

### Covariates

Models were adjusted for age (continuous), sex, race/ethnicity (non-Hispanic White, non-Hispanic Black, Mexican American, other Hispanic, other race), education (less than high school, high school/GED, some college, college graduate or above), body mass index (continuous, kg/m^2^), smoking status (never, former, current), self-reported physician-diagnosed diabetes (yes/no), and hypertension (yes/no).

### Statistical Analysis

Baseline characteristics were compared across RHR clinical categories (<60, 60–69, 70–79, ≥80 bpm) using Kruskal-Wallis tests for continuous variables and chi-square tests for categorical variables. Crude mortality rates were calculated per 1000 person-years.

The primary analysis used Cox proportional hazards regression with follow-up time (years from examination to death or 31 December 2019) as the time scale. Models were built sequentially: Model 1 (unadjusted), Model 2 (age, sex, race/ethnicity), and Model 3 (fully adjusted for all covariates). The dose–response relationship was examined using restricted cubic splines with 4 knots placed at the 5th, 35th, 65th, and 95th percentiles. Inverse probability weighted (IPW) Kaplan-Meier survival curves were constructed for RHR clinical categories and tertiles, with weights derived from multinomial propensity score models adjusting for all covariates. Weighted likelihood ratio test P values were used for group comparisons. To illustrate the non-cardiovascular pathway, all Kaplan-Meier and dose–response analyses were presented side by side for all-cause and non-CVD mortality, with cardiovascular deaths treated as censored events.

Cause-specific Cox models were fitted for CVD mortality, cancer mortality, accidental death, and non-CVD mortality, with deaths from other causes treated as censored events. Subgroup analyses were conducted by age group (20–29, 30–39, 40–49), sex, race/ethnicity, smoking status, BMI category, and hypertension status; interaction P values were obtained from likelihood ratio tests of cross-product terms. Fine-grained age-stratified analyses used 5-year age bands.

Sensitivity analyses included exclusion of deaths within 2 and 5 years of baseline, use of an alternative RHR measurement (treadmill warm-up heart rate), additional adjustment for physical activity and laboratory values, survey-weighted Cox regression incorporating NHANES complex survey design, and substitution of individual comorbidities (diabetes, hypertension) with a modified Charlson Comorbidity Index constructed from available NHANES questionnaire data.^19^

Model discrimination was assessed using Harrell’s C-statistic, and internal validation was performed using 200 bootstrap resamples. Predicted 15-year mortality probabilities were estimated from the fully adjusted Cox model for representative demographic profiles to illustrate risk heterogeneity.

All analyses were conducted in R version 4.5.2. Two-sided P < .05 was considered statistically significant.

## Results

### Study Population

The analytic sample comprised 3291 participants (mean [SD] age, 33.6 [8.3] years; 52% male) with a mean (SD) RHR of 71.2 (10.7) bpm (Table 1; eFigure 1). Those with higher RHR tended to be younger, were more likely to smoke, and had higher BMI (eTable 1). Over a median follow-up of 17.8 years, 120 deaths occurred.

| Characteristic | Resting Heart Rate Category |  |  |  |  | p-value<sup>2</sup> |
| --- | --- | --- | --- | --- | --- | --- |
|  | Overall N =<br>3,359<sup>1</sup> | <60 bpm N =<br>421<sup>1</sup> | 60–69 bpm N =<br>1,028<sup>1</sup> | 70–79 bpm N =<br>1,112<sup>1</sup> | ≥80 bpm N =<br>798<sup>1</sup> |  |
| Age (years) | 33.2 (8.6) | 34.1 (8.4) | 33.9 (8.6) | 33.0 (8.6) | 32.4 (8.5) | <0.001 |
| Sex |  |  |  |  |  | <0.001 |
| Female | 1,615 (48.1%) | 105 (24.9%) | 452 (44.0%) | 583 (52.4%) | 475 (59.5%) |  |
| Male | 1,744 (51.9%) | 316 (75.1%) | 576 (56.0%) | 529 (47.6%) | 323 (40.5%) |  |
| Race/Ethnicity |  |  |  |  |  | 0.075 |
| Mexican American | 854 (25.4%) | 108 (25.7%) | 284 (27.6%) | 274 (24.6%) | 188 (23.6%) |  |
| Non-Hispanic Black | 659 (19.6%) | 95 (22.6%) | 203 (19.7%) | 203 (18.3%) | 158 (19.8%) |  |
| Non-Hispanic White | 1,571 (46.8%) | 194 (46.1%) | 466 (45.3%) | 527 (47.4%) | 384 (48.1%) |  |
| Other Hispanic | 162 (4.8%) | 19 (4.5%) | 46 (4.5%) | 57 (5.1%) | 40 (5.0%) |  |
| Other Race | 113 (3.4%) | 5 (1.2%) | 29 (2.8%) | 51 (4.6%) | 28 (3.5%) |  |
| Education |  |  |  |  |  | 0.10 |
| Less than high school | 761 (22.7%) | 89 (21.2%) | 246 (23.9%) | 250 (22.5%) | 176 (22.1%) |  |
| High school / GED | 806 (24.0%) | 88 (21.0%) | 247 (24.0%) | 263 (23.7%) | 208 (26.1%) |  |
| Some college | 1,052 (31.3%) | 132 (31.4%) | 306 (29.8%) | 348 (31.3%) | 266 (33.4%) |  |
| College graduate or above | 738 (22.0%) | 111 (26.4%) | 229 (22.3%) | 251 (22.6%) | 147 (18.4%) |  |
| Unknown | 2 | 1 | 0 | 0 | 1 |  |
| BMI (kg/m<sup>2</sup>) | 27.4 (5.9) | 26.3 (4.6) | 26.9 (5.3) | 27.7 (6.0) | 28.3 (6.9) | <0.001 |
| Unknown | 10 | 0 | 2 | 2 | 6 |  |
| Waist circumference (cm) | 92.5 (14.4) | 90.1 (12.1) | 91.3 (13.1) | 93.1 (15.0) | 94.2 (16.0) | <0.001 |
| Unknown | 26 | 2 | 7 | 7 | 10 |  |
| Smoking status |  |  |  |  |  | <0.001 |
| Current | 890 (26.5%) | 110 (26.2%) | 257 (25.0%) | 271 (24.4%) | 252 (31.7%) |  |
| Former | 530 (15.8%) | 82 (19.5%) | 187 (18.2%) | 155 (14.0%) | 106 (13.3%) |  |
| Never | 1,935 (57.7%) | 228 (54.3%) | 584 (56.8%) | 685 (61.7%) | 438 (55.0%) |  |
| Unknown | 4 | 1 | 0 | 1 | 2 |  |
| Alcohol use |  |  |  |  |  | 0.054 |
| Drinker | 788 (75.8%) | 109 (83.8%) | 245 (77.5%) | 240 (73.6%) | 194 (72.4%) |  |
| Never | 252 (24.2%) | 21 (16.2%) | 71 (22.5%) | 86 (26.4%) | 74 (27.6%) |  |
| Unknown | 2,319 | 291 | 712 | 786 | 530 |  |
| Diabetes | 54 (1.6%) | 3 (0.7%) | 5 (0.5%) | 19 (1.7%) | 27 (3.4%) | <0.001 |
| Unknown | 1 | 0 | 0 | 0 | 1 |  |
| Hypertension | 294 (8.9%) | 22 (5.3%) | 84 (8.3%) | 97 (8.9%) | 91 (11.6%) | 0.003 |
| Unknown | 51 | 4 | 19 | 16 | 12 |  |
|  | Overall N =<br>3,359<sup>1</sup> | <60 bpm N =<br>421<sup>1</sup> | 60–69 bpm N =<br>1,028<sup>1</sup> | 70–79 bpm N =<br>1,112<sup>1</sup> | ≥80 bpm N =<br>798<sup>1</sup> |  |
| Resting heart rate (bpm) | 71.3 (10.7) | 54.8 (3.6) | 64.4 (2.8) | 73.7 (2.8) | 85.8 (5.4) | <0.001 |
| Peak heart rate (bpm) | 141.8 (17.1) | 139.6 (16.6) | 139.7 (18.3) | 142.4 (16.7) | 144.7 (15.9) | <0.001 |
| Absolute HRR (bpm) | 70.4 (19.2) | 84.9 (16.8) | 75.3 (18.7) | 68.7 (16.8) | 58.9 (16.6) | <0.001 |
| % age-predicted HR achieved | 75.9 (8.1) | 75.1 (7.8) | 75.0 (8.7) | 76.1 (8.0) | 77.1 (7.6) | <0.001 |
| Chronotropic index | 0.6 (0.1) | 0.6 (0.1) | 0.6 (0.1) | 0.6 (0.1) | 0.6 (0.1) | <0.001 |
| HR recovery at 1 min (bpm) | 10.6 (7.6) | 13.9 (8.0) | 11.7 (8.2) | 10.3 (6.5) | 7.9 (7.1) | <0.001 |
| Unknown | 359 | 55 | 88 | 137 | 79 |  |
| HR recovery at 2 min (bpm) | 28.0 (11.7) | 35.0 (11.8) | 30.3 (11.2) | 27.4 (10.9) | 22.4 (10.7) | <0.001 |
| Unknown | 266 | 34 | 75 | 98 | 59 |  |
| Estimated VO<sub>2</sub>max (ml/kg/min) | 40.1 (10.0) | 45.2 (10.4) | 41.3 (9.7) | 38.9 (9.7) | 37.4 (9.3) | <0.001 |
| Unknown | 57 | 2 | 16 | 23 | 16 |  |
| HbA1c (%) | 5.3 (0.7) | 5.2 (0.4) | 5.2 (0.5) | 5.3 (0.6) | 5.4 (1.0) | 0.13 |
| Unknown | 112 | 13 | 32 | 38 | 29 |  |
| Total cholesterol (mg/dL) | 192.6 (39.1) | 190.7 (38.7) | 189.9 (37.6) | 193.5 (39.1) | 195.9 (40.8) | 0.011 |
| Unknown | 143 | 18 | 44 | 43 | 38 |  |
| C-reactive protein (mg/dL) | 0.3 (0.7) | 0.2 (0.3) | 0.3 (0.4) | 0.4 (0.7) | 0.5 (0.9) | <0.001 |
| Unknown | 128 | 16 | 39 | 40 | 33 |  |
| Follow-up (years) | 17.6 (2.2) | 17.6 (2.3) | 17.7 (2.2) | 17.7 (2.2) | 17.5 (2.2) | 0.027 |
| Unknown | 3 | 0 | 3 | 0 | 0 |  |
| All-cause death | 124 (3.7%) | 13 (3.1%) | 32 (3.1%) | 43 (3.9%) | 36 (4.5%) | 0.4 |
| CVD death | 24 (0.7%) | 2 (0.5%) | 8 (0.8%) | 7 (0.6%) | 7 (0.9%) | 0.8 |
| Cancer death | 42 (1.3%) | 7 (1.7%) | 12 (1.2%) | 10 (0.9%) | 13 (1.6%) | 0.4 |

<sup>1</sup>Mean (SD); n (%)
<sup>2</sup>Kruskal-Wallis rank sum test; Pearson's Chi-squared test

### Cause-of-Death Distribution

The mortality profile differed markedly from that of older populations. Cancer was the leading cause of death (42 [35.0%]), followed by NCHS residual causes (31 [25.8%]), a category encompassing suicide, chronic liver disease, HIV, and other conditions. Accidents accounted for 20 deaths (16.7%), with the remaining non-CVD deaths attributed to chronic lower respiratory disease (3), diabetes (2), and nephritis (1). Cardiovascular disease accounted for 21 deaths (17.5%; heart disease, 17; cerebrovascular disease, 4). Overall, non-CVD causes accounted for 99 of 120 deaths (82.5%; eTable 2).

### RHR and All-Cause Mortality

In the fully adjusted model, each 10-bpm increase in RHR was associated with 26% higher all-cause mortality (HR, 1.26; 95% CI, 1.07–1.50; P = .007; Table 2). IPW-adjusted Kaplan-Meier curves showed visible survival divergence across RHR categories (weighted LRT P < .001; Figure 1, Panels A–B), and the association followed a monotonic gradient across RHR quintiles (Q5 vs Q1: HR, 1.95; 95% CI, 1.07–3.55; P for trend = .014; eTable 3). Restricted cubic spline analysis confirmed a linear dose–response relationship (P for nonlinearity = .86; Figure 2, Panel A).

**Table 2.**
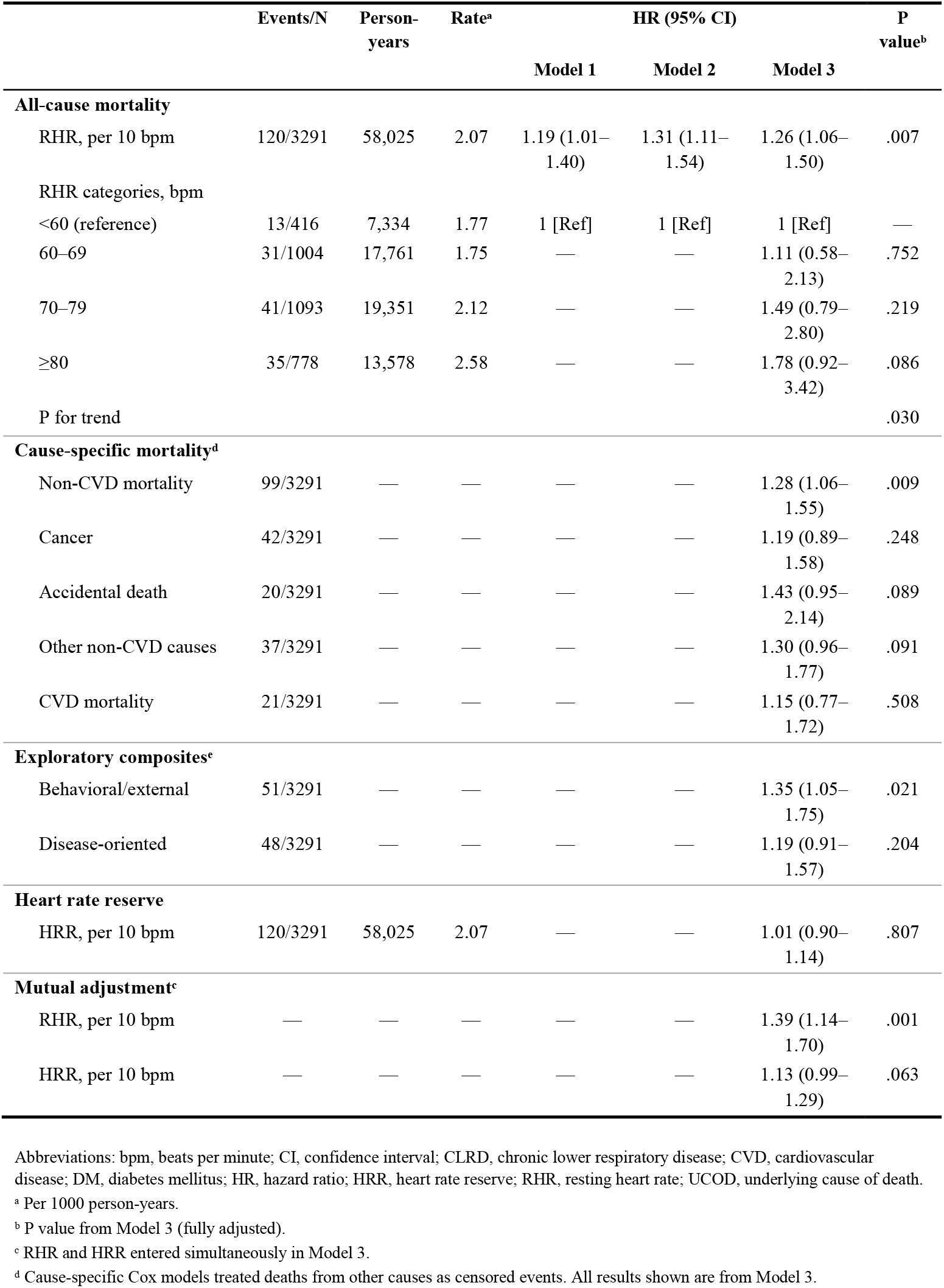
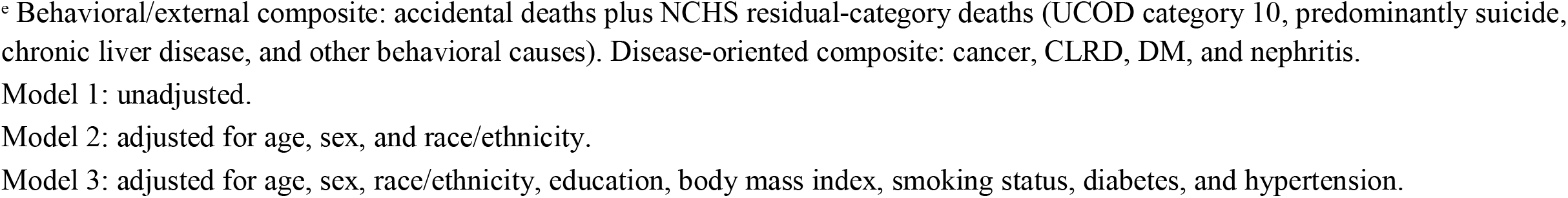
Association of Resting Heart Rate and Heart Rate Reserve With All-Cause and Cause-Specific Mortality Among US Adults Aged 20 to 49 Years, NHANES 1999–2004.

### RHR and Cause-Specific Mortality

The central finding was a dissociation between cardiovascular and non-cardiovascular pathways (Figure 4). RHR was significantly associated with non-CVD mortality (HR, 1.28; 95% CI, 1.07– 1.55; P = .009) but not with CVD mortality (HR, 1.15; 95% CI, 0.77–1.71; P = .51).

To localize the non-CVD signal, we constructed two exploratory composites. A behavioral/external composite (accidental deaths plus NCHS residual-category deaths; 51 events) was significantly associated with RHR (HR, 1.35; 95% CI, 1.05–1.75; P = .02), whereas a disease-oriented composite (cancer, chronic lower respiratory disease, diabetes, and nephritis; 48 events) was not (HR, 1.19; P = .20; eTable 5). This divergence suggests that the non-CVD mortality signal is concentrated among causes linked to behavioral risk and autonomic dysregulation rather than chronic organ disease. Individual non-CVD subcategories showed consistently elevated but individually nonsignificant hazard ratios: accidental death (HR, 1.42; P = .09), other non-CVD causes (HR, 1.30; P = .09), and cancer (HR, 1.19; P = .25). Among the 111 deaths without diabetes or hypertension on the death certificate, the RHR association remained significant (HR, 1.26; 95% CI, 1.05–1.50; P = .01).

When restricted to participants aged 35 years and older, the RHR association with non-CVD mortality strengthened considerably (57 events; HR, 1.52; 95% CI, 1.19–1.94; P < .001), and cancer-specific mortality also became significant (28 events; HR, 1.50; 95% CI, 1.05–2.13; P = .03; Figure 4).

The IPW-adjusted Kaplan-Meier curves for non-CVD mortality showed more pronounced separation than those for all-cause mortality (Figure 1, Panels B and D vs A and C), visually confirming that the survival disadvantage of elevated RHR operates through non-cardiovascular pathways (eFigure 4). This pattern was consistent in dose–response analyses, which showed significant linear associations for RHR with both all-cause (P-linear = .007) and non-CVD mortality (P-linear = .009), with no evidence of nonlinearity (Figure 2, Panels A–B).

### Age-Dependent Emergence

The RHR–mortality association was age dependent. Among participants younger than 35 years, no association was observed (HR, 0.98; 95% CI, 0.74–1.30; P = .88), whereas among those aged 35 to 49 years, the association was strong (HR, 1.48; 95% CI, 1.19–1.83; P < .001). Analysis by 5-year age bands localized the inflection point to ages 35 to 39 (HR, 2.60; 95% CI, 1.46–4.64; P = .001; eFigure 5), with sustained significance at ages 40 to 44 (HR, 1.43; 95% CI, 1.02–2.01; P = .038; eTable 10).

### Heart Rate Reserve

Heart rate reserve was not associated with all-cause mortality (HR, 1.01; 95% CI, 0.90–1.14; P =.81; eTable 4) or any cause-specific outcome (all P > .18). RHR and HRR were weakly correlated (eFigures 2–3). This null finding was consistent across all age strata, sex, race/ethnicity, and comorbidity subgroups (Figure 3; eTable 9).

### Risk Heterogeneity Across Demographic Profiles

To translate relative hazard ratios into clinically interpretable absolute risks, we estimated 15-year all-cause mortality from the fully adjusted Cox model for 30 demographic profiles defined by age, sex, and race/ethnicity, all at RHR = 80 bpm (Figure 5; eFigure 6).

Age was the dominant modifier. Among non-Hispanic White females, predicted mortality rose steadily from 1.1% at age 25 to 1.4% at age 30, 1.7% at age 35, 2.1% at age 40, and 2.6% at age 45. The same age gradient was observed across all sex and race/ethnicity strata, consistent with the age-dependent emergence of the RHR association.

Sex approximately doubled absolute risk at every age. At age 35, predicted mortality was 1.7% for White females versus 3.3% for White males; at age 45, it was 2.6% versus 5.0%.

Race/ethnicity introduced additional heterogeneity within each age-sex stratum. Among males at age 45, predicted mortality was 4.7% for Mexican Americans, 5.0% for non-Hispanic Whites, and 6.9% for non-Hispanic Blacks. Among females at age 45, corresponding values were 2.4%, 2.6%, and 3.5%.

The cumulative effect of these three factors produced a more than 6-fold gradient at a single RHR value: from 1.0% (25-year-old Mexican American female) to 6.9% (45-year-old non-Hispanic Black male). These results indicate that a fixed RHR threshold cannot capture individual-level risk and that clinical interpretation of elevated RHR requires consideration of the full demographic context.

### Sensitivity Analyses

The RHR–all-cause mortality association was robust across multiple sensitivity analyses: exclusion of early deaths (≤2 years: HR, 1.24, P = .014; ≤5 years: HR, 1.29, P = .007), use of an alternative RHR measurement (HR, 1.38, P < .001), and additional adjustment for physical activity (HR, 1.27, P = .006) and laboratory values (HR, 1.29, P = .006; eTable 8). Substituting a modified Charlson Comorbidity Index for individual comorbidities yielded virtually identical results (HR, 1.28; P = .005; eTable 8d). Survey-weighted analysis was directionally consistent, though attenuated (HR, 1.18; 95% CI, 0.99–1.42; eTable 7). The E-value for unmeasured confounding was 1.84 (lower CI bound, 1.26). Model discrimination statistics are reported in eTable 6. Bootstrap internal validation showed modest optimism (apparent C-statistic, 0.710; corrected, 0.682; eTable 8b).

## Discussion

In this nationally representative cohort of 3291 US adults aged 20 to 49 years followed for a median of 17.8 years, we observed three findings that, to our knowledge, have not been previously reported in this population. First, consistent with prior observations,^29^ elevated RHR predicted all-cause mortality, but the association appeared to be driven by non-cardiovascular rather than cardiovascular causes of death. Second, this association was absent before age 35 and emerged thereafter, suggesting a risk window below the starting age of current screening tools.

Third, heart rate reserve under submaximal conditions showed no prognostic value across any outcome or subgroup.

### A Non-Cardiovascular Pathway

RHR predicted non-CVD mortality (HR, 1.28; P = .009) but not CVD mortality (HR, 1.15; P =.51) in this young adult cohort (Table 2). This finding extends the observation by Greenland et al^29^ that RHR predicts noncardiovascular mortality in younger men. Our analysis adds to this evidence by showing that, in this cohort, the observed mortality signal was driven by non-cardiovascular pathways, with no statistically significant association for cardiovascular mortality. These data suggest a reappraisal of the prevailing framework linking elevated RHR to mortality primarily through atherosclerotic cardiovascular disease.^5,6^ In older populations, the RHR–mortality association is largely mediated by coronary artery disease, heart failure, and sudden cardiac death.^1–4^ In young adults, whose atherosclerotic burden is minimal, elevated RHR may instead mark a constellation of non-cardiovascular risk factors.

Exploratory cause-specific analyses showed that the non-CVD mortality signal was not driven by any single cause of death. The hazard ratios for accidental death (HR, 1.42; P = .09), other non-CVD causes (HR, 1.30; P = .09), and cancer (HR, 1.19; P = .25) were uniformly elevated but individually underpowered. Combining accidental deaths and NCHS residual-category deaths into a behavioral/external composite yielded a significant association (51 events; HR, 1.35; 95% CI, 1.05–1.75; P = .02), whereas a disease-oriented composite (cancer, chronic lower respiratory disease, diabetes, and nephritis; 48 events) did not (HR, 1.19; P = .20). This divergent pattern suggests that the non-CVD mortality signal is concentrated among causes linked to behavioral risk rather than chronic organ disease.

The profile of residual-category decedents reinforced this interpretation. Among the 31 deaths classified as NCHS category 10 (“all other causes”), none had baseline diabetes, only 6.5% had hypertension, and 93.5% had neither condition on the death certificate. Based on national vital statistics, NCHS category 10 in younger age groups is predominantly composed of suicide, homicide, and chronic liver disease/cirrhosis, which together account for an estimated 65% to 80% of this residual category.^17,18^ These causes share a common upstream pathway: autonomic dysregulation characterized by sympathetic overactivation, the same physiological state indexed by elevated RHR.

Several biological and behavioral mechanisms connect sympathetic overactivation to these specific causes of death. Elevated RHR reflects chronic sympathetic nervous system activation, which is associated with psychological distress, impulsivity, and substance use disorders, conditions disproportionately prevalent in young adults and strongly linked to suicide, homicide, and alcohol-related liver disease.^10,11,22^ The borderline association with accidental death (HR, 1.42; P = .09), the majority of which among younger decedents reflects drug overdose and motor vehicle incidents,^23^ is also consistent with evidence linking autonomic dysregulation to risk-taking behavior.^12^ Additionally, elevated RHR is a marker of chronic low-grade inflammation, which may contribute to cancer mortality through non-atherosclerotic pathways. The RHR– cancer association became significant only when restricted to participants aged 35 years and older (HR, 1.50; 95% CI, 1.05–2.13; P = .03), consistent with the latency required for inflammation-driven carcinogenesis.^13,14^

### Age-Dependent Emergence

The RHR–mortality association was absent before age 35 but emerged sharply thereafter, with the steepest inflection at ages 35 to 39. This pattern likely reflects the accumulation of subclinical organ damage that transforms elevated RHR from a transient physiological state into a clinically significant risk marker.^20^ This threshold falls below the starting age of current risk stratification tools (SCORE2 begins at age 40; the Pooled Cohort Equations at age 40), suggesting a potential gap in preventive screening for adults in their mid-thirties.^15,16^

### HRR Under Submaximal Conditions

The uniformly null finding for HRR carries practical implications. Heart rate reserve measured at submaximal intensity (75% of age-predicted maximum) may lack the physiological stress necessary to unmask chronotropic incompetence, which has been linked to mortality in studies using maximal exercise testing.^9,21^ Population-based surveys and primary care settings that rely on submaximal protocols should therefore interpret HRR results with caution.

### Clinical Implications

These findings suggest that elevated RHR in young adults should not be interpreted solely through a cardiovascular lens. Clinicians encountering persistently elevated RHR in patients younger than 50 years may consider evaluating for underlying contributors such as psychological distress, substance use, sleep disorders, or chronic inflammation, rather than focusing exclusively on cardiovascular risk factor modification.

Predicted 15-year mortality ranged from 1% to 7% across demographic profiles at RHR = 80 bpm, highlighting the need for individualized risk communication. Although the relative hazard associated with RHR did not differ significantly by sex or race/ethnicity (all P-interaction > .67; eTable 9), the absolute risk gradient across these groups was not merely a statistical artifact. Young males die disproportionately from accidents, suicide, homicide, and substance-related causes,^24^ the same behavioral/external category that drove the RHR–mortality association in this study. Non-Hispanic Black individuals face a disproportionate burden of chronic psychosocial stress, a well-established driver of sustained sympathetic activation and accelerated physiological aging.^25,26^ In both cases, the reason these groups carry higher baseline mortality overlaps substantially with the mechanism through which RHR predicts death: chronic sympathetic overactivation fueling behavioral and external-cause mortality. RHR may therefore capture autonomic vulnerability operating at both the individual and population levels.

### Limitations

Several limitations merit discussion. First, the 120 deaths (21 CVD) limited statistical power for individual cause-specific estimates. However, the primary contrast between CVD and non-CVD mortality was adequately powered, and the composite reclassification analyses provided a coherent pattern consistent with the behavioral/external pathway hypothesis. Second, NHANES public-use mortality files classify cause of death into 10 broad categories (UCOD_LEADING), and the residual category 10 cannot be disaggregated into individual causes (eg, suicide, homicide, chronic liver disease) without restricted-use data. We addressed this by referencing national vital statistics distributions for younger age groups to characterize category 10 composition.^17,18^ Third, RHR was measured once at baseline; however, single-visit RHR has demonstrated acceptable reproducibility in epidemiological studies,^27^ and any regression dilution would bias results toward the null, making our estimates conservative.^28^ Fourth, residual confounding by unmeasured factors (eg, mental health status, substance use, sleep quality) is possible in any observational study. The E-value of 1.84 (lower bound, 1.26) indicates that a moderately strong unmeasured confounder would be needed to fully explain the observed association. We further addressed confounding through multiple sensitivity analyses: substituting a modified Charlson Comorbidity Index for individual comorbidities yielded virtually identical results (HR, 1.26-1.28; all P < .01; eTable 8d),^19^ and restricting to deaths without diabetes or hypertension on the death certificate produced consistent findings (111 events; HR, 1.26; P =.01). Fifth, these findings require replication in independent cohorts with detailed cause-of-death coding to confirm the pathway specificity observed here.

## Conclusions

In this nationally representative cohort of US adults aged 20 to 49 years followed for nearly two decades, elevated RHR predicted all-cause mortality through non-cardiovascular rather than cardiovascular pathways, with the signal concentrated among behavioral and external causes of death. The association was absent before age 35 and emerged sharply thereafter, below the starting age of current cardiovascular screening tools. Heart rate reserve measured under submaximal conditions carried no prognostic value. These results suggest that RHR in young adults may serve as a marker of global health vulnerability rather than cardiovascular risk alone, and that clinical evaluation of persistently elevated RHR in this age group may benefit from considering psychological distress, substance use, and chronic inflammation alongside traditional cardiovascular risk factors.

## Data Availability

The NHANES data used in this study are publicly available from the National Center for Health Statistics (https://www.cdc.gov/nchs/nhanes/). The linked mortality files are available from the NCHS (https://www.cdc.gov/nchs/data-linkage/mortality-public.htm). Complete analysis code is available upon reasonable request to the corresponding author.

https://www.cdc.gov/nchs/nhanes/

---

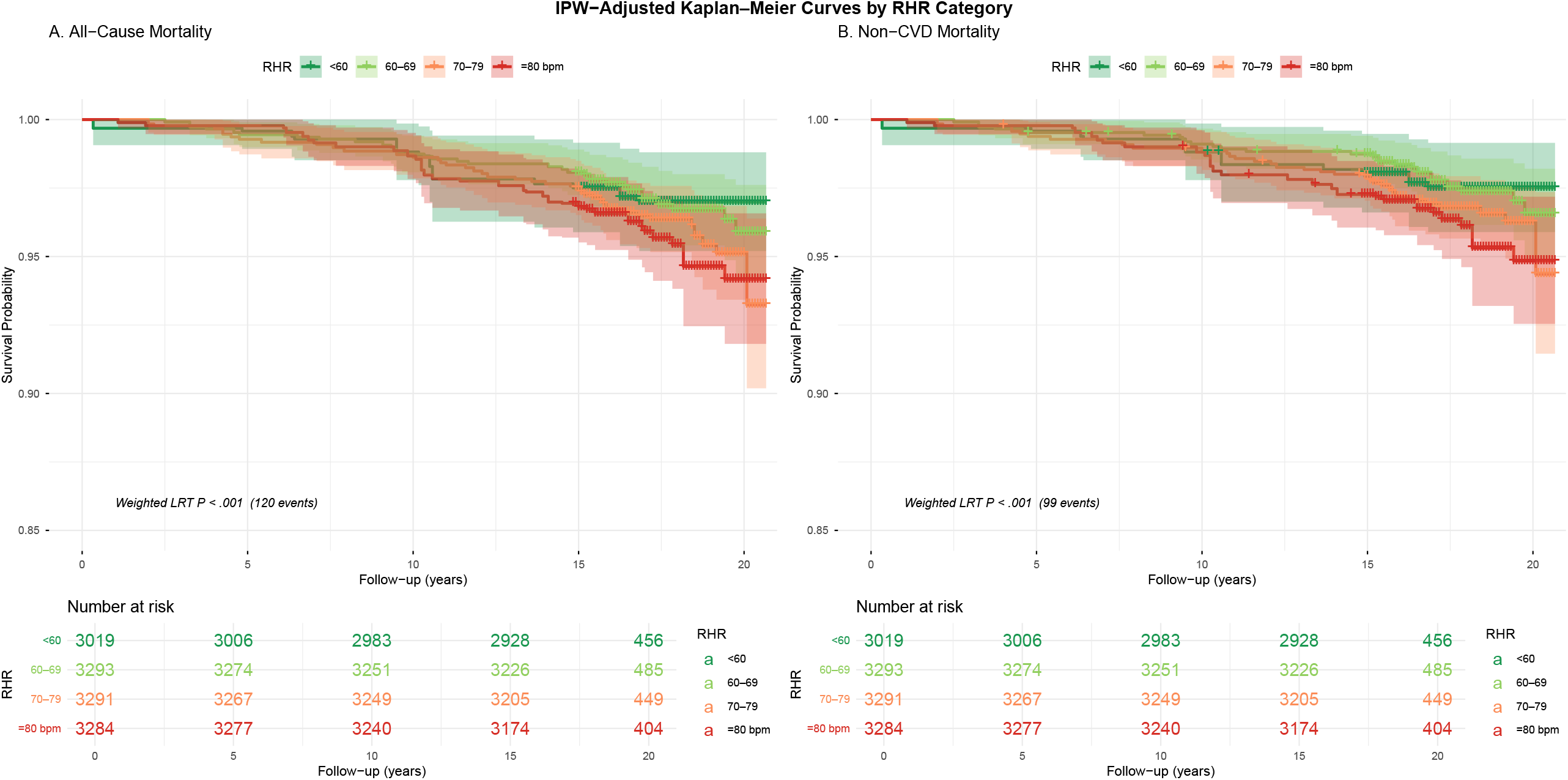

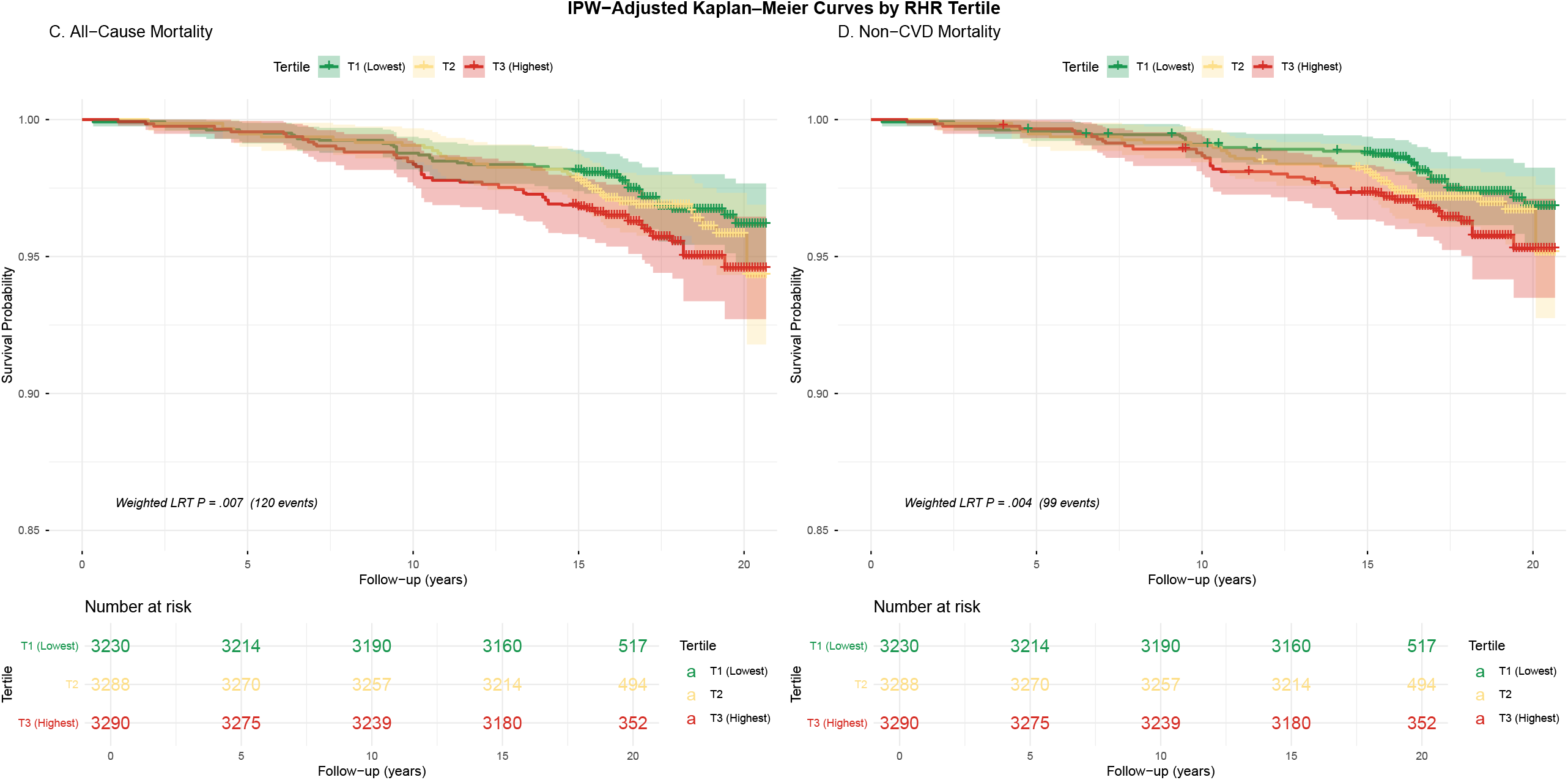

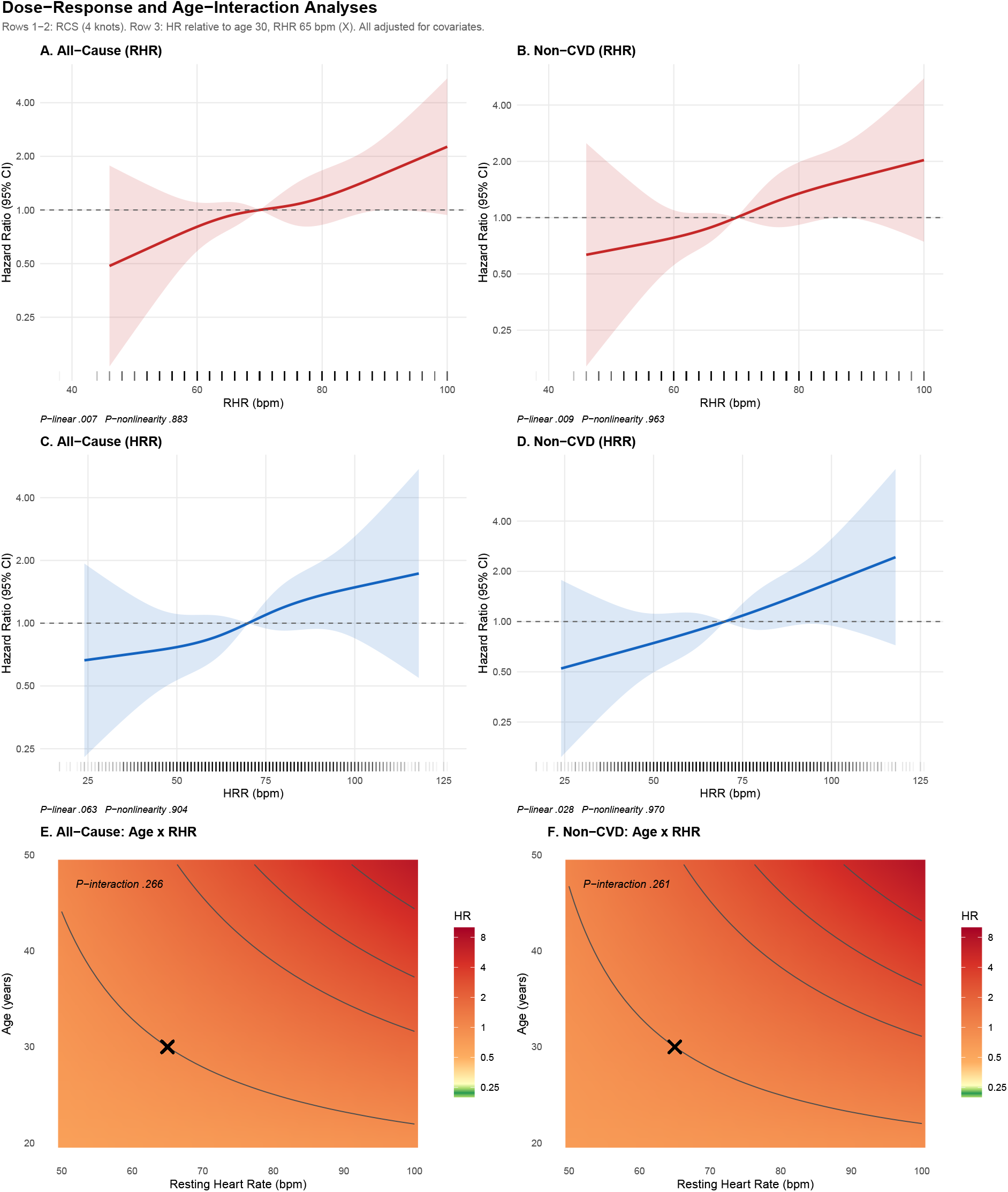

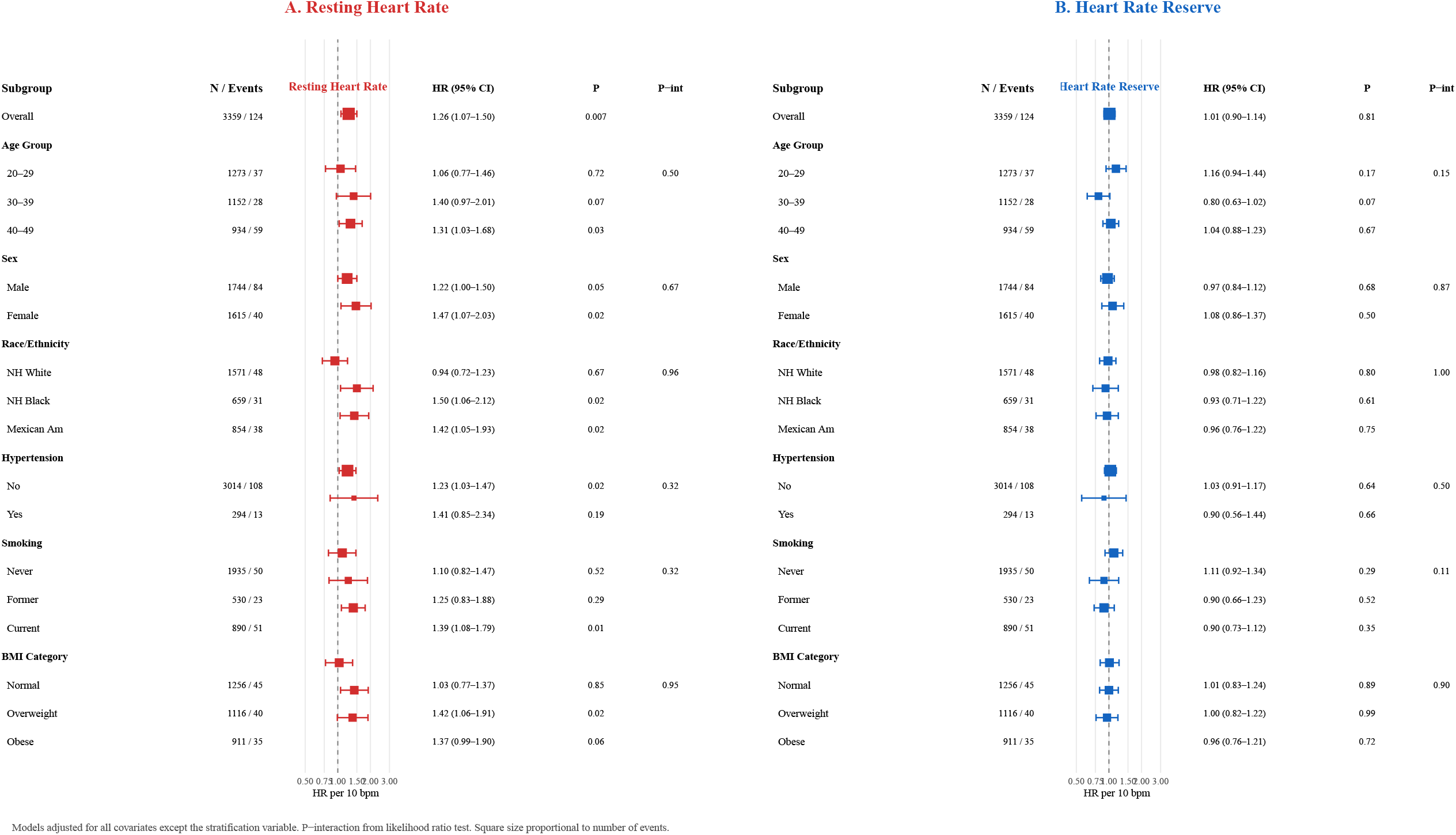

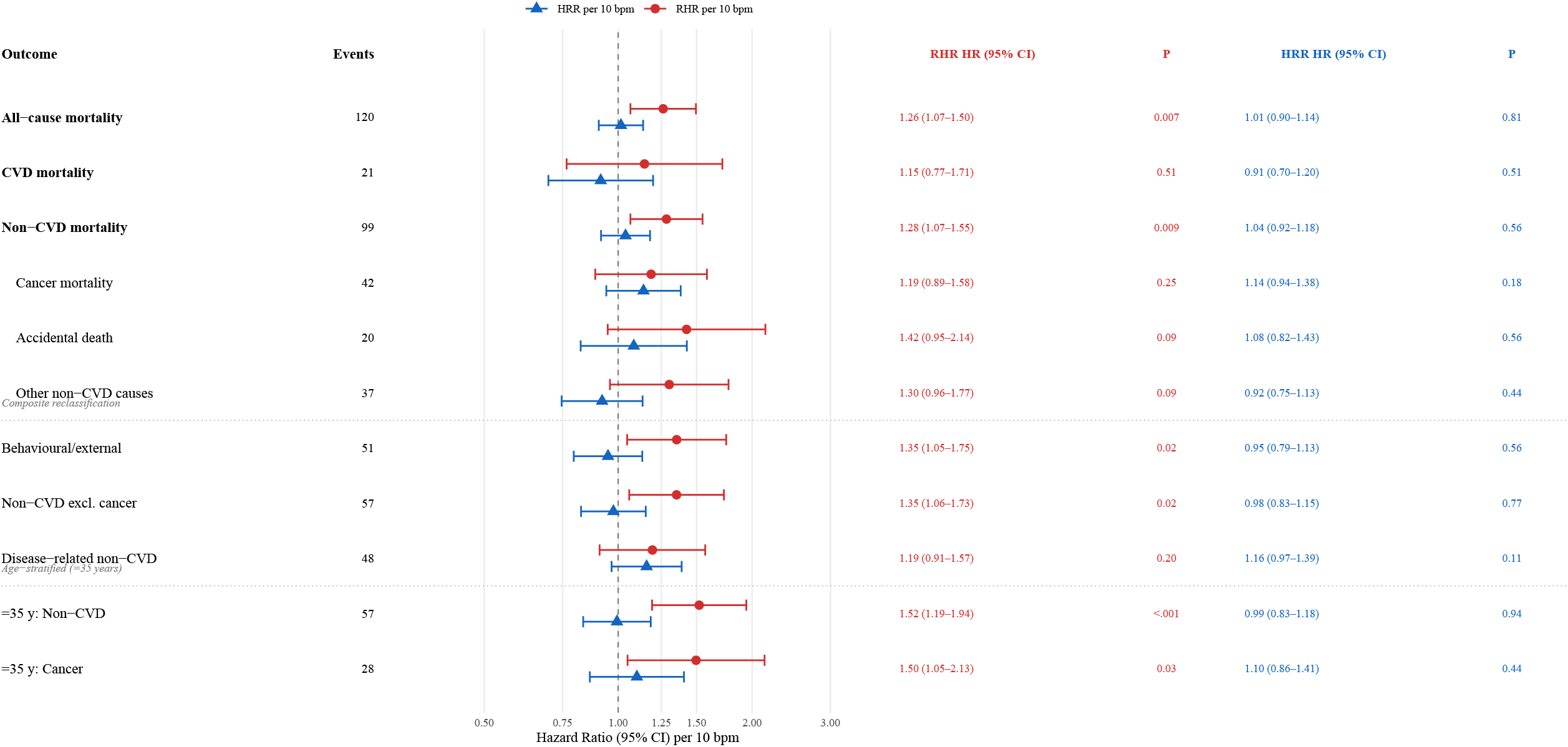

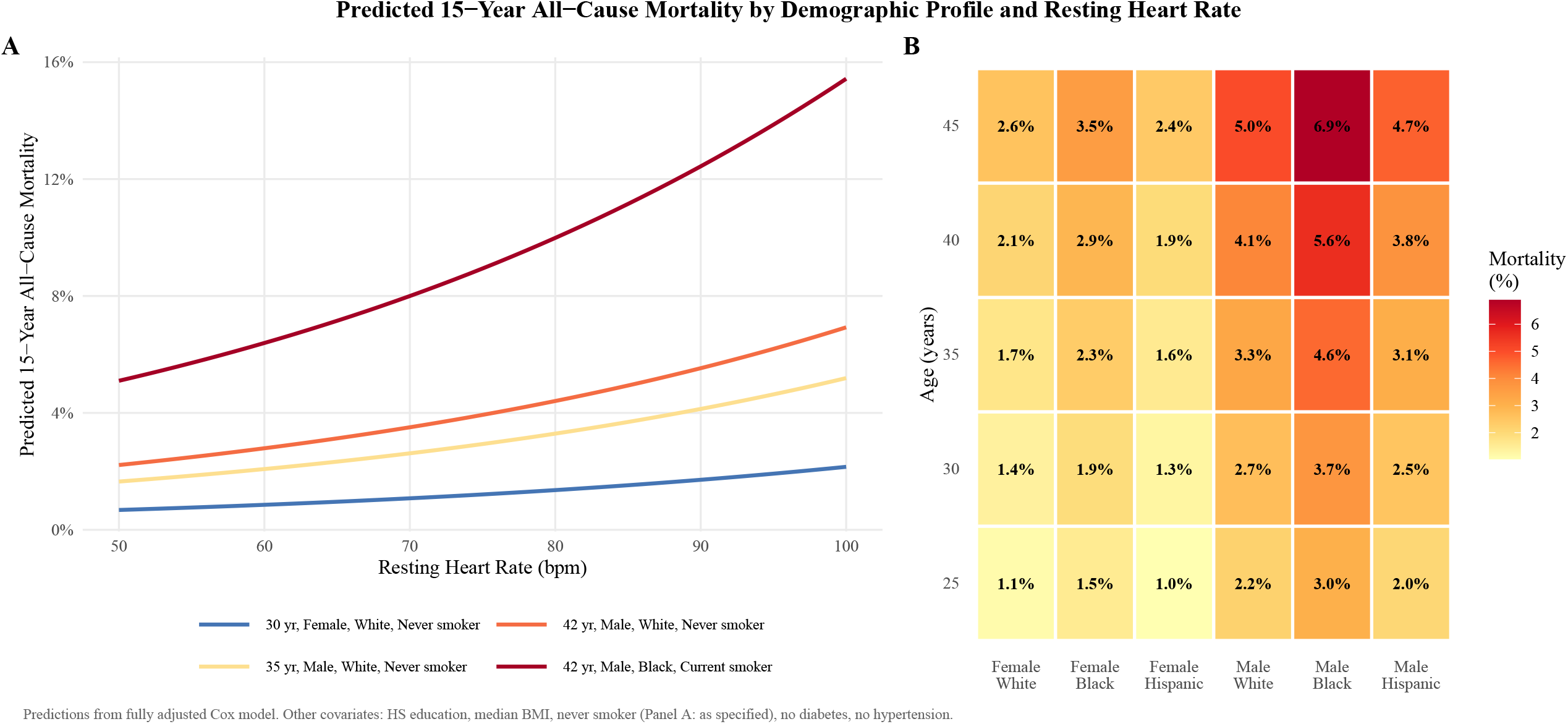

